# Association of state social distancing restrictions with nursing home COVID-19 and non-COVID-19 outcomes

**DOI:** 10.1101/2021.02.08.21251382

**Authors:** Yue Li, Zijing Cheng, Xueya Cai, Yunjiao Mao, Helena Temkin-Greener

## Abstract

The COVID-19 poses a disproportionate threat to nursing home residents. Although recent studies suggested the effectiveness of state social distancing measures in the United States on curbing COVID-19 morbidity and mortality among the general population, there is lack of evidence as to how these state orders may have affected nursing home patients or what potential negative health consequences they may have had. In this longitudinal study, we evaluated changes in state strength of social distancing restrictions from June to August of 2020, and their associations with the weekly numbers of new COVID-19 cases, new COVID-19 deaths, and new non–COVID-19 deaths in nursing homes of the US. We found that stronger state social distancing measures were associated with improved COVID-19 outcomes (case and death rates), reduced across-facility disparities in COVID-19 outcomes, but more deaths due to non–COVID-19 reasons among nursing home residents.

## INTRODUCTION

The coronavirus disease 2019 (COVID-19) poses a disproportionate threat to older adults especially those residing in long-term care (LTC) facilities such as nursing homes or assisted living facilities.^1-5^ COVID-19 is known to have been present in almost all of the more than 15,500 nursing homes in the United States. By December 23, 2020, over 905,000 coronavirus cases and 113,000 associated deaths were reported in LTC facilities, representing 6% and 39% of total COVID-19 cases and deaths, respectively.^6^

Recent studies suggest that racial/ethnic disparities in COVID-19 morbidity and mortality among LTC residents^2,4,7,8^ largely mirror the disparities found in the general population.^9-12^ In a national study of nursing homes, Li and colleagues revealed that COVID-19 cases and deaths per facility were 2 to 4 times higher in nursing homes with highest proportions of racial/ethnic minority residents than in nursing homes with low proportions.^7^ These disparities are largely a result of system-level inequalities and segregation of care in that older residents of color are disproportionately cared for in facilities that are located in marginalized communities, have inadequate resources and limited abilities to respond to outbreaks of emerging infections, and provide care of poorer quality.^13^

During the pandemic, deaths from non–COVID-19 causes, such as Alzheimer disease, diabetes, and heart disease, also increased markedly, mostly among older adults.^14,15^ Delay or avoidance of necessary medical care because of concerns about COVID-19 and social distancing restrictions increases morbidity and mortality risk associated with the health conditions of older adults, and may contribute to reported excess deaths.^16,17^ Moreover, disruptions of in-person social activities during the pandemic have led to increase in social isolation and loneliness among older adults, another serious public health issue that is associated with psychological suffering, unmet personal and health care needs, and deaths.^18-21^

Starting from early March, most states in the US implemented non-pharmaceutical public health interventions to contain coronavirus transmission. These policies took a variety of forms – such as statewide shelter-in-place orders, closure of non-essential businesses, and bans on large and small-group gatherings – and were implemented at different times and with different levels of enforcement across states.^22^ Recent studies suggested the effectiveness of these state social distancing measures on curbing COVID-19 cases, hospitalizations, and associated deaths among the general population.^22-26^ However, there is lack of evidence as to how these state orders may have affected the most vulnerable groups, such as LTC patients, or what potential negative health consequences they may have had, such as excess mortality due to reasons other than COVID-19 infection.

In this longitudinal study, we evaluated changes in state strength of social distancing restrictions from June to August of 2020, and their associations with the weekly numbers of new COVID-19 cases, new COVID-19 deaths, and new non–COVID-19 deaths in nursing homes nationally. We hypothesized that stronger state COVID-19 restrictions led to reduced COVID-19 case and death rates, reduced disparities in COVID-19 case and death rates between nursing homes with high and low proportions of racial/ethnic minorities, but increased non–COVID-19 mortality rate among nursing home residents.

## RESULTS

### Descriptive characteristics of the sample

Table 1 presents that among the 14,046 nursing homes that submitted the COVID-19 data for the week of August 17-23 (one week after state COVID-19 restrictions were ranked on August 11), 6,829 nursing homes (48.6%) were in states with low COVID-19 restrictions and 7,217 (51.4%) were in states with high restrictions. Nursing homes in states with high COVID-19 restrictions tended to be larger, for-profit facilities with slightly better nurse staffing and five-star ratings. They also tended to have higher cumulative numbers of COVID-19 cases (among residents and staff) and deaths (among residents) before state social distancing restrictions were ranked, and to be located in larger counties with more COVID-19 cases and deaths as well as states with higher COVID-19 death rates. Supplementary Table S1 shows that nursing homes with higher proportions of racial/ethnic minority residents tended to be larger, for-profit, and chain facilities that serve a higher proportion of Medicaid residents, have lower nurse staffing and five-star ratings, and are located in larger counties with more COVID-19 cases and deaths.

**Table 1.** Nursing home, county, and state characteristics by strength of state social distancing measures reported on August 11, 2020.

| Nursing home characteristic | All nursing homes, counties, or states | Strength of state social distancing measures* |  |
| --- | --- | --- | --- |
|  |  | Low | High |
|  |  | Mean±SD or N (%) |  |
| Number of nursing homes | 14,046 | 6,829 (48.62) | 7,217 (51.38) |
| Weekly number of new Covid-19 confirmed cases among residents<sup>a</sup> | 0.59±2.80 | 0.70±2.94 | 0.49±2.66 |
| 0 | 12,142 (86.44) | 5,745 (84.13) | 6,397 (88.64) |
| 1-10 | 1,532 (10.91) | 890 (13.03) | 642 (8.90) |
| >10 | 372 (2.64) | 194 (2.84) | 178 (2.47) |
| Weekly number of new Covid-19 confirmed cases among staff<sup>a</sup> | 0.51±1.63 | 0.59±1.72 | 0.42±1.52 |
| 0 | 10,981 (78.18) | 5,151 (75.43) | 5,830 (80.78) |
| 1-10 | 2,835 (20.18) | 1,562 (22.87) | 1,273 (17.64) |
| >10 | 230 (1.64) | 116 (1.70) | 114 (1.58) |
| Weekly number of new Covid-19 related deaths among residents<sup>b</sup> | 0.10±0.55 | 0.12±0.60 | 0.08±0.49 |
| 0 | 13,125 (93.44) | 6,316 (92.49) | 6,809 (94.35) |
| 1-5 | 744 (5.30) | 429 (6.28) | 315 (4.36) |
| >5 | 177 (1.26) | 84 (1.23) | 93 (1.29) |
| Weekly number of new non-Covid-19 related deaths among residents<sup>b</sup> | 0.41±1.84 | 0.38±1.37 | 0.45±2.20 |
| 0 | 10,382 (73.92) | 5,073 (74.30) | 5,309 (73.56) |
| 1-5 | 3,463 (24.66) | 1,667 (24.41) | 1,796 (24.89) |
| >5 | 200 (1.42) | 88 (1.29) | 112 (1.55) |
| Total number of certified beds | 106.24±55.86 | 101.23±49.96 | 110.98±60.54 |
| Number of residents | 85.69±48.48 | 79.15±41.75 | 91.89±53.35 |
| Ownership |  |  |  |
| For-profit | 9,862 (70.21) | 4,632 (67.83) | 5,230 (72.47) |
| Non-profit | 3,295 (23.46) | 1,677 (24.56) | 1,618 (22.42) |
| Government owned | 889 (6.33) | 520 (7.61) | 369 (5.11) |
| Chain affiliated | 8,272 (58.89) | 4,042 (59.19) | 4,230 (58.89) |
| Hospital affiliated | 541 (3.85) | 230 (3.37) | 311 (4.31) |
| Percentage of Medicaid residents, % | 59.84±23.08 | 59.65±22.40 | 60.03±23.70 |
| Percentage of Medicare residents, % | 13.80±13.31 | 13.20±13.09 | 14.37±13.48 |
| Case mix index score | 1.29±0.16 | 1.26±0.14 | 1.31±0.18 |
| RN hours per resident day | 0.67±0.48 | 0.64±0.44 | 0.70±0.51 |
| Total nurse hours per resident day | 3.83±0.88 | 3.76±0.82 | 3.89±0.93 |
| Overall five-star rating | 3.19±1.41 | 3.14±1.40 | 3.24±1.42 |
| Cumulative number of Covid-19 confirmed cases among residents before August 11<sup>c</sup> | 2.68±8.81 | 2.18±8.06 | 3.16±9.44 |
| Cumulative number of Covid-19 confirmed cases among staff before August 11<sup>c</sup> | 2.46±7.11 | 2.07±7.77 | 2.84±6.41 |
| Cumulative number of Covid-19 deaths among residents before August 11<sup>c</sup> | 0.76±2.82 | 0.52±2.34 | 0.98±3.20 |
| **County characteristic** |  |  |  |
| Cumulative number of Covid-19 cases before August 11, x1k<sup>c</sup> | 3.67±12.47 | 1.58±4.55 | 5.68±16.65 |
| Cumulative number of Covid-19 deaths before August 11, x1k<sup>c</sup> | 0.09±0.26 | 0.04±0.13 | 0.15±0.34 |
| Total populationx100k | 8.25±17.94 | 4.35±9.35 | 11.93±22.72 |
| **State characteristic** |  |  |  |
| Cumulative rate of Covid-19 cases before August 11 (per one thousand)<sup>c</sup> | 3.56±3.11 | 3.82±3.31 | 3.30±2.88 |
| Cumulative rate of Covid-19 deaths before August 11 (per one thousand)<sup>c</sup> | 0.10±0.08 | 0.08±0.05 | 0.13±0.10 |
| Percentage population ≥65 years, % | 14.86±2.00 | 15.19±1.73 | 14.56±2.19 |
| Percentage of non-white population, % | 30.40±10.78 | 29.26±10.45 | 31.49±10.98 |

\* All p-values were <0.001 for comparisons of group differences based on t-tests for continuous variables and chi-square tests for categorical variables, except for the following characteristics: weekly number of new non-Covid-19 related deaths among residents, three categories (p=0.320); chain affiliated (p=0.487); hospital affiliated (p=0.004); and percentage of Medicaid residents (p=0.326).
<sup>a</sup> Numbers are for the reporting week ending on August 23 (Monday August 17 to Sunday August 23).
<sup>b</sup> Numbers are for the reporting week ending on September 6 (Monday August 31 to Sunday September 6).
<sup>c</sup> Cumulative numbers are reported for the period from May 25 to August 9, 2020.
SD=standard deviation; RN=registered nurse.

### Association of state COVID-19 restrictions with nursing home COVID-19 outcomes

For the week of August 17-23, and compared to nursing homes in states with low COVID-19 restrictions, nursing homes in states with high COVID-19 restrictions had fewer new COVID-19 confirmed cases among residents (0.49 vs 0.70 per facility on average) and among staff (0.42 vs 0.59 per facility on average), as well as fewer new COVID-19 related deaths among residents (0.08 vs 0.12 per facility on average; Table 1). Similar differences were found for other study weeks (results not shown).

Multivariable longitudinal analyses (Table 2) confirmed that nursing homes in states with stronger social distancing restrictions had both the reduced likelihood of having at least one weekly reported new case (or death) and, among nursing homes with ≥1 case (or death), fewer new cases (or deaths). For example, high (vs low) state COVID-19 restrictions was independently associated with a 19% reduced likelihood (odds ratio [OR]=0.81, 95% confidence interval [CI] 0.76-0.87, p<0.001) of having ≥1 new resident case and among nursing homes with ≥1 case, a 15% reduction (incidence rate ratio [IRR]=0.85, 95% CI 0.79-0.91, p<0.001) in new cases. On average, higher stringency of state social distancing measures helped to reduce weekly new cases among residents by 0.13 cases/facility (p<0.001; from 0.67 to 0.54 cases/facility) as shown in Figure 1, panel A. Sensitivity analyses confirmed the robustness of these results when state policy rankings were specified as a continuous variable or categorized as tertiles (Supplementary Tables S2 and S3).

**Table 2.**
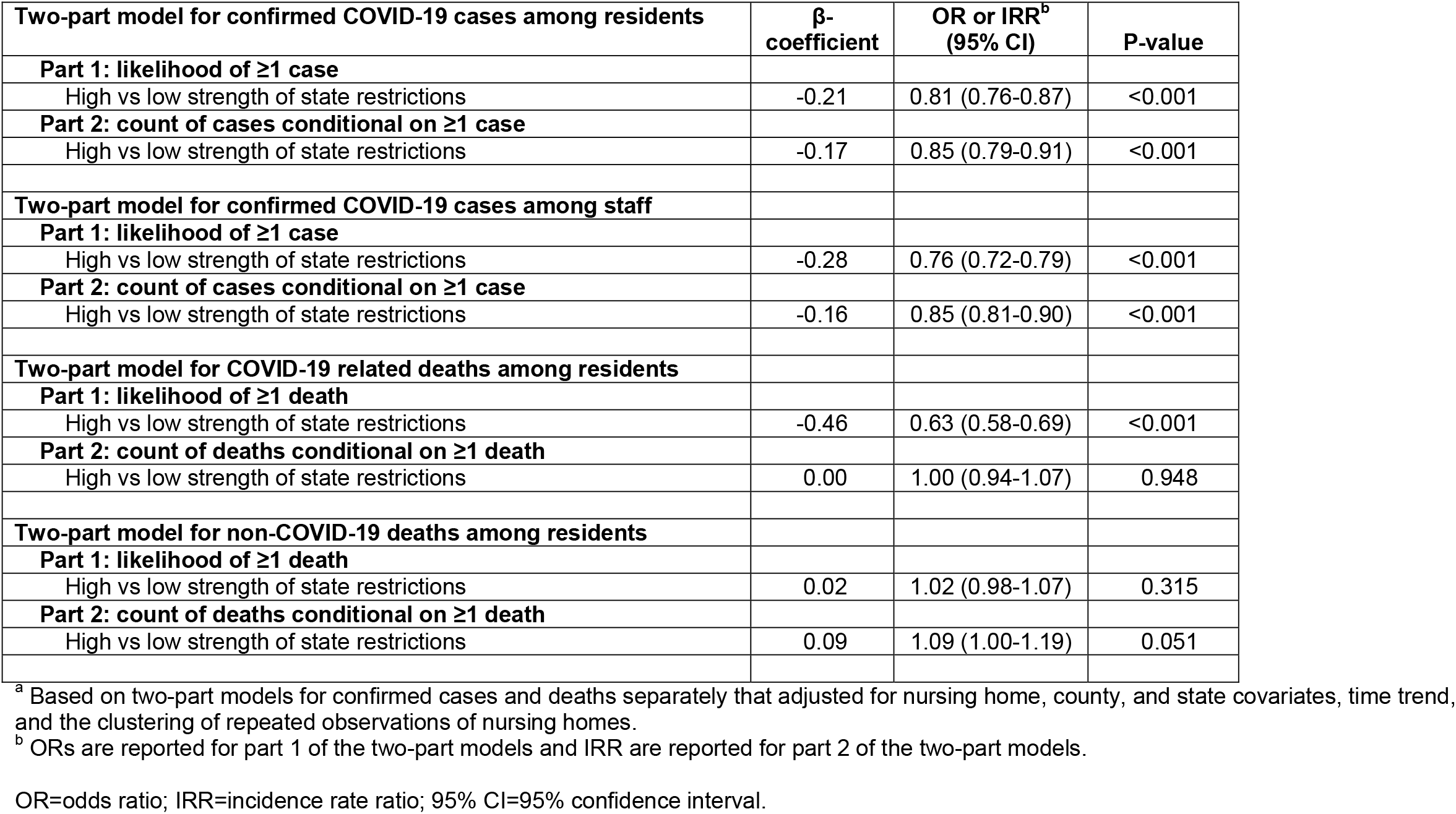
Associations between the strength of state social distancing measures and nursing home COVID-19 and non-COVID-19 outcomes.^a^.

**Figure 1.**
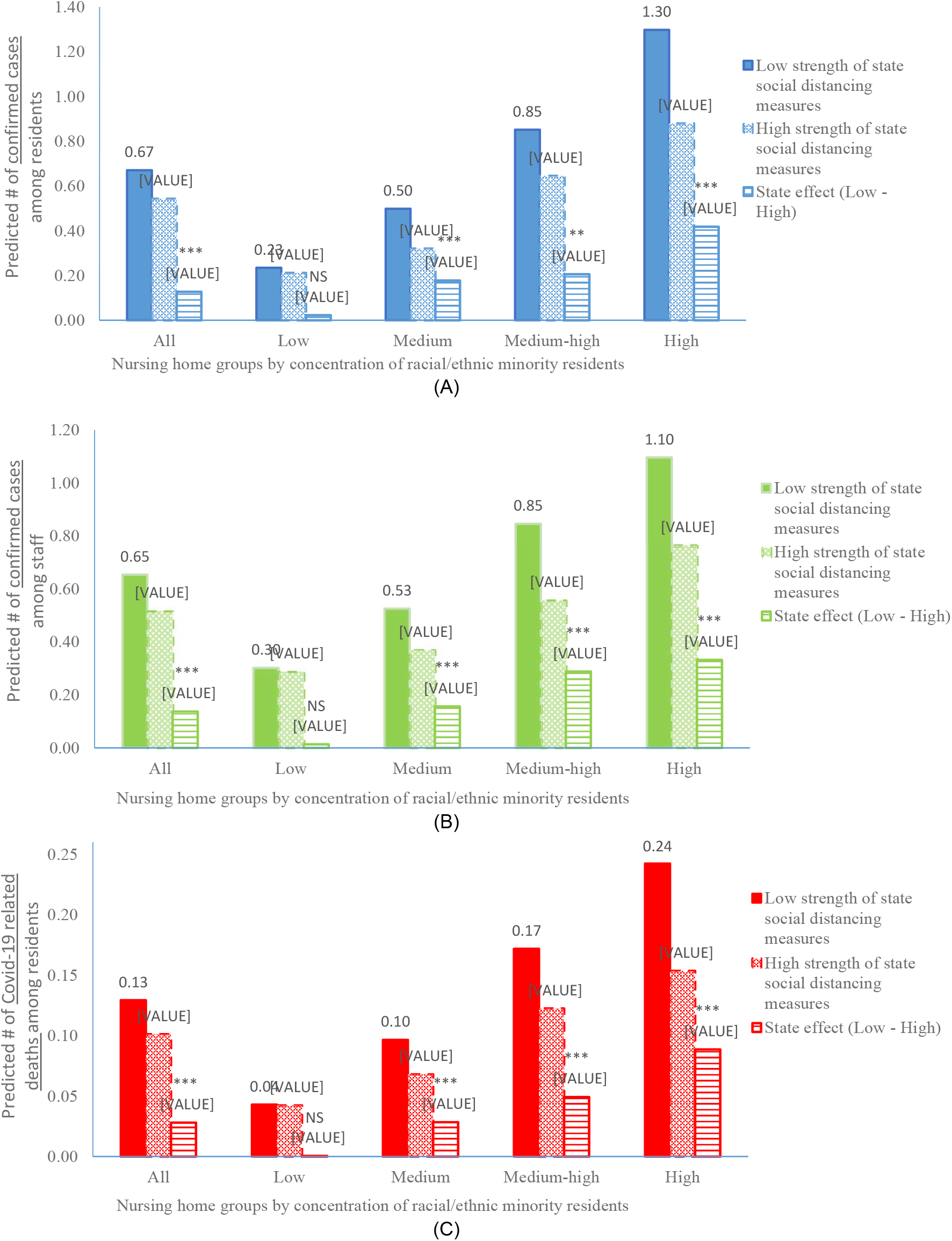
Predicted numbers of (A) weekly new laboratory-confirmed COVID-19 cases among residents, (B) new weekly laboratory-confirmed COVID-19 cases among staff, and (C) new weekly COVID-19 related deaths among residents reported in U.S. nursing homes for the week of July 5, 2020 (Monday June 29 to Sunday July 5) to the week of September 6, 2020 (Monday August 31 to Sunday September 6), by strength of state social distancing measures (high vs low). Predicted numbers are presented for all nursing homes and by nursing home groups with different percentages of racial and ethnic minority residents. Note: *p<0.05, **p<0.01, ***p<0.001, and NS=not significant (p≥0.05). P-values indicate the statistical significance of reductions in predicted numbers between nursing homes in states with high strength of social distancing measures and nursing homes in states with low social distancing measures, and were derived from the joint tests of the two-part regression models. Two-part models were used to derive the predicted numbers and their differences due to state policy effect. All two-part models adjusted for nursing home, county, and state covariates, time trend, and the clustering of repeated observations of nursing homes.

### Association of state COVID-19 restrictions with disparities in nursing home COVID-19 outcomes

Existing evidence suggests across-facility disparities in COVID-19 case and death rates.^2,4,7,8^ Table 3 shows that stronger state COVID-19 restrictions may help reduce such disparities by reducing the likelihood of having 1 or more new cases (or deaths) in a reporting week and reducing the number of cases (or deaths) for nursing home serving disproportionately more racial/ethnic minority residents. For example, although high (vs low) state COVID-19 restrictions was not associated with the likelihood of having ≥1 case (OR=1.05, 95% CI 0.89-1.25, p=0.536) or the conditional count of cases (IRR=0.85, 95% CI 0.70-1.04, p=0.112) among residents for nursing homes with low concentrations of minority residents, the corresponding associations (OR=0.72, 95% CI 0.65-0.80, p<0.001; and IRR=0.78, 95% CI 0.70-0.87, p<0.001) were evident for nursing homes with high concentrations of minority residents, leading to a 31% (95% CI −17% to −44%, p<0.001) reduced disparity in the likelihood of having ≥1 case between the two groups, although the reduced disparity in conditional count (9%) were not statistically significant (95% CI −27% to 14%, p=0.435). In Figure 1, we similarly show stronger state policy effect on reduced numbers of cases and deaths among nursing homes with a higher proportion of racial/ethnic minority residents.

**Table 3.**
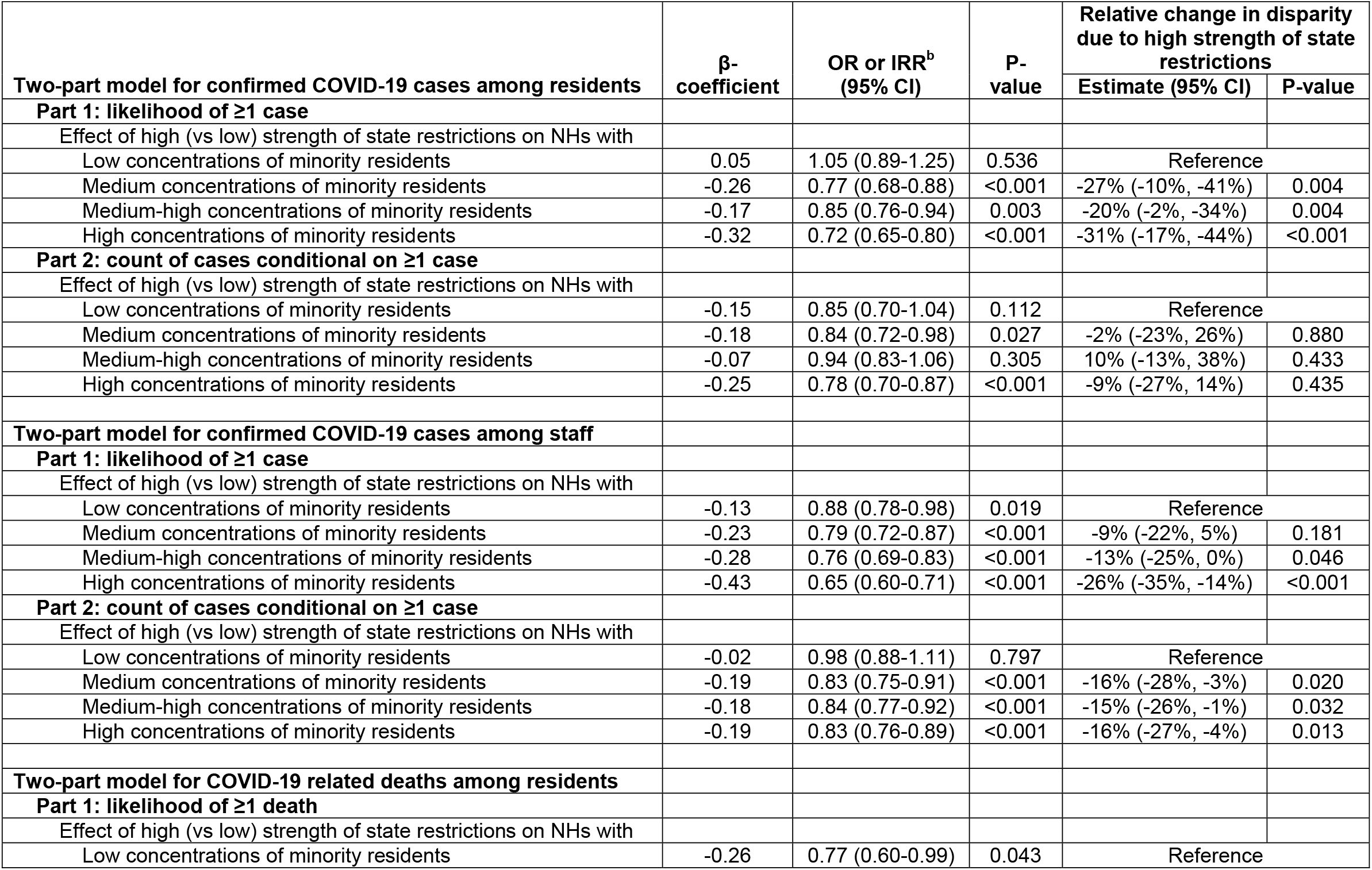
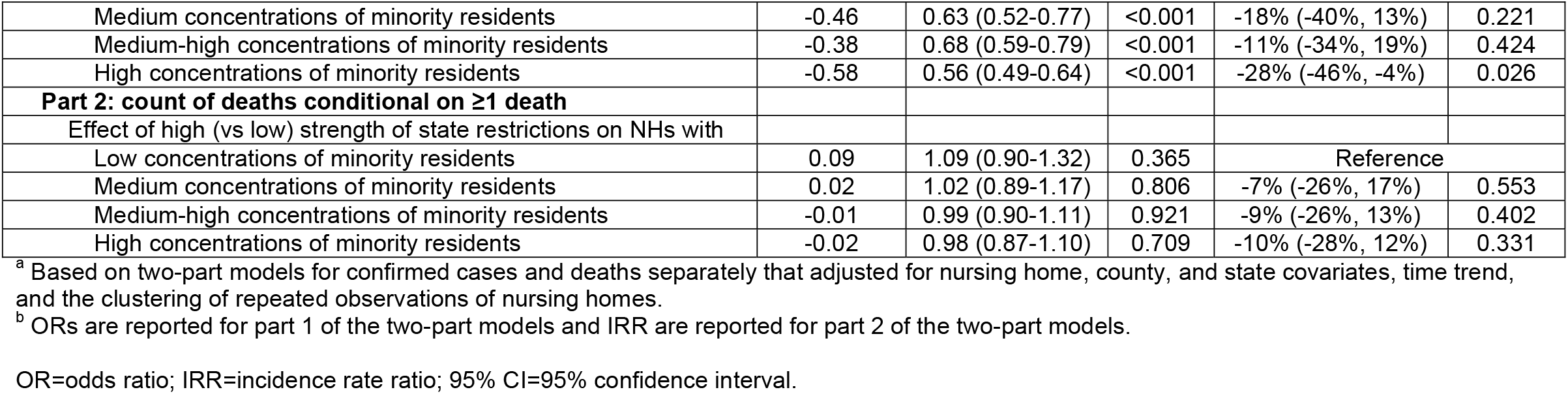
Associations between the strength of state social distancing measures and disparities in COVID-19 outcomes across nursing homes (NHs) serving different proportions of racial/ethnic minority residents.^a^.

### Association of state COVID-19 restrictions with non–COVID-19 deaths among residents

For the week of August 17-23, non-COVID-19 death rates were higher in nursing homes in states with low compared to those in states with high COVID-19 restrictions (0.45 vs 0.38 per facility on average; Table 1). Similar differences were found for other study weeks (results not shown).

Multivariable longitudinal analyses (Table 2) confirmed that high (vs low) state COVID-19 restrictions was not independently associated with the likelihood of having ≥1 new non-COVID-19 death (OR=1.02, 95% CI 0.98-1.07, p=0.315); however, it was independently associated with a 9% increase in non-COVID-19 death count (IRR=1.09, 95% CI 1.00-1.19, p=0.051) among nursing homes with ≥1 new non-COVID-19 death. The two-part model predicted that high strength of state social distancing measures on average led to increased weekly non–COVID-19 death rate among residents by 0.10 cases/facility (0.41 and 0.51 deaths/facility for nursing homes in states with low and high COVID-19 restrictions, respectively; p=0.090 for difference).

Sensitivity analyses did not suggest an association between state COVID-19 restrictions and non-COVID-19 deaths when state rankings were included in the model as a continuous variable (Supplementary Table S2), but suggested such an association (with marginal statistical significance) when state rankings were categorized as tertiles (Supplementary Table S3). Compared to nursing homes in the 1^st^ tertile group, nursing homes in the 3^rd^ tertile group has an OR of 0.95 (95% CI 0.90-1.02, p=0.139) for the likelihood of having ≥1 non-COVID-19 death and an IRR of 1.10 (95% CI 0.98-1.23, p=0.105) for conditional count of non-COVID-19 deaths. The predicted difference in overall count of non-COVID-19 deaths between the two groups was 0.10 (0.39 and 0.49 deaths/facility for nursing homes in the 1^st^ and the 3^rd^ tertile group, respectively; p=0.090 for difference).

## DISCUSSION

This study found that stronger state social distancing measures were associated with lower weekly rates of new COVID-19 confirmed cases and related deaths among nursing home residents, as well as lower weekly COVID-19 new confirmed case rate among nursing home staff, from June to September of 2020. The magnitude of these associations was larger for nursing homes serving disproportionately more racial/ethnic minority residents, suggesting that stronger state social distancing measures also helped reduce disparities in COVID-19 outcomes between nursing homes with higher and lower proportions of minority residents as reported in recent studies. Our analyses also found that stronger state social distancing measures were associated with a somewhat increased rate of non-COVID-19 mortality among nursing home residents, suggesting a potential unintended consequence of restrictions on in-person social activities on excess mortality.

Since early March and in the wake of the novel coronavirus outbreak at a skilled nursing facility in Washington state,^1^ the CMS, in coordination with state agencies and the Centers for Disease Control and Prevention (CDC), has immediately refocused their quality of care inspections in nursing homes on compliance with infection control policies and practices.^27^ The CMS and the CDC also released guidelines and rules to combat the surge of COVID-19 infections and deaths in LTC facilities such as restrictions on nursing home visitors including non-essential health care personnel;^28^ cancellation of all group activities and communal dining in nursing homes; implementation of active screening of residents and health care personnel for fever and respiratory symptoms;^29^ and a mandate to separate staff members dedicated to the care of residents with COVID-19 from those caring for residents without (or whose COVID-19 status is unknown).^30^

In addition to these federal guidelines specifically designed to curb transmission of the virus in LTC facilities, most states in the US implemented social distancing measures to contain the growth rate of infections among the general population following the experiences of countries attacked earlier by the pandemic.^22-24^ The details of these social distancing restrictions, and their implementation or enforcement, vary over states and over time as the pandemic progresses; for example, many states partially reopened non-essential businesses in early summer but closed these businesses again due to surges of COVID-19 in the second wave of the pandemic. Although recent studies examined variations of these state social distancing measures and demonstrated their effectiveness on reducing COVID-19 related morbidity and mortality,^22-26^ the state COVID-19 policies examined in these studies were generally limited to the early stage of the pandemic (i.e. March to May of 2020). It thus remains uncertain if the effect of strong state COVID-19 restrictions has been maintained as the pandemic continues to evolve, and if prolonged restrictions may lead to serious negative health consequences such as excess mortality.

These expected state policy effects, positive or negative, would likely manifest first and strongest among nursing home residents who represent the most vulnerable group and who are hit the hardest throughout different stages of the pandemic. However, thus far, there has been a lack of studies to determine the effect of early state (and federal) responses to the pandemic on COVID-19 outcomes among nursing home residents, largely due to the absence of nursing home COVID-19 data nationally during early period of the pandemic.

In an effort to fill this information gap, the CMS released guidance on April 19 that required nursing homes to report COVID-19 cases and deaths directly to the CDC,^31^ and a later interim final rule of the CMS on May 1 marked the formal start of this national data submission process. These data were first publicly published on June 4, containing facility-level counts of COVID-19 cases and deaths since late May; they continue to be updated weekly by the CMS. Taking advantage of this new data reporting system and state variations in the restrictiveness of their social distancing policies, this study found stronger state COVID-19 restrictions were associated with reduced nursing home COVID-19 infection and fatality rates, results that one would expect to see. Of note, this study was not able to determine the effect of above-mentioned federal responses to the COVID-19 outbreaks in LTC facilities. These federal guidelines and rules likely helped avoid a large number of COVID-19 cases and deaths in all nursing homes in the US, but their exact effects are difficult, if not impossible, to determine due to the lack of a comparison group (e.g. nursing homes not subjecting to these federal rules). The variation in state COVID-19 restrictions provide a natural experiment for investigating the effect of state policies. In addition, several features of this study, such as its longitudinal design and the control of the secular trend and the cumulative number of COVID-19 cases (or deaths) right before state COVID-19 policies were ranked in multivariable analyses, allowed us to estimate the independent effect of state COVID-19 policies that is above and beyond the effect of federal guidelines and rules designed to curb coronavirus transmissions in all nursing homes.

The stronger effect of state COVID-19 policies on improved COVID-19 outcomes for nursing homes that disproportionately care for more racial/ethnic minority residents suggests an additional disparity reduction effect of these state policies. Since the beginning of the global COVID-19 pandemic in March, compelling evidence has indicated that racial/ethnic minority persons are at higher risk of COVID-19 infections, hospitalizations, and deaths, compared to non-Hispanic whites.^9-12^ A most recent study of the Kaiser Family Foundation^12^ has analyzed electronic health records of 50 million patients and estimated that although there is little difference in the testing rates by race and ethnicity, Black, Hispanic, and Asian patients are about two-to-three times as likely to test positive as Whites. Moreover, hospitalization and death rates are at least twice as high among people of color as among White patients, and these disparities persist after accounting for differences in sociodemographic characteristics and underlying health conditions.

Racial/ethnic disparities in outcomes of nursing home care have existed long before the COVID-19 pandemic,^13,32,33^ largely due to system-level inequalities and segregation of care. These disparities persisted in the past several decades despite evidence of overall improved quality of care and outcomes for all residents and nursing homes over time. The across-facility disparities in COVID-19 morbidity and mortality reported by recent studies^2,7,8^ are just another manifestation of the structural inequalities in nursing home care that have existed for decades. For example, nursing homes serving disproportionately non-white residents tend to be faced with serious resource constrains and operate within local healthcare systems that are also resource strained and thus easily overwhelmed by the pandemic.

The Coronavirus Aid, Relief, and Economic Security (CARES) Act enacted on March 27, 2020 intended to blunt the impact of economic downturn due to pandemic through financial aid to businesses, individuals, and healthcare institutions.^34^ Under the Provider Relief Fund authorized by the CARES Act, the federal government has allocated $10 billion to nursing homes thus far which helped all facilities in the nation to address shortages in personal protective equipment (PPE) and staff and to improve testing capacities.^35^ Nevertheless, these federal funds are “color blind” and do not explicitly target systemic inequalities in nursing home care under the COVID-19 pandemic.^36^ It is unknown if and how this federal aid may help to address the across-facility disparities in COVID-19 outcomes.

Our findings of the reduced disparities in nursing home COVID-19 outcomes as a result of stronger state social distancing stipulations suggest the possibility that broadly targeted policy and public health interventions are able to both mitigate coronavirus transmissions and redress outcome disparities due to enduring system-level inequalities. Although the exact mechanism through which disparities are reduced is not clear, it appears that certain level of stringency in social distancing restrictions are particularly necessary for the subgroups of minority-serving and resource poor nursing homes to effectively reduce virus transmissions among their residents and staff. It is also possible that in states with low strength of COVID-19 restrictions, the weekly COVID-19 case and mortality rates for nursing homes caring for low proportions of racial/ethnic minority residents were already very low (0.23/facility and 0.04/facility, respectively; Figure 1) compared to nursing homes caring for high proportions of such residents (1.30/facility and 0.24/facility, respectively). Thus, stronger state social distancing policies may not be able to further reduce case and mortality rates for the former group of nursing homes due to floor effects.

Concerns about the downside effect of prolonged social distancing restrictions have been expressed, although empirical evidence is limited on how these restrictions affect the physical and mental health outcomes of individuals.^14-17,21^ Deaths in the US attributed to noninfectious causes, such as heart disease and dementia, increased throughout the spring and summer surges in COVID-19 cases,^14,15^ possibly due to disruptions in medical care access and delivery subsequent to shelter-in-place orders. It is conceivable that compared to community-living older adults, nursing home residents are more vulnerable to disrupted care routines given their advanced ages, complex morbidity patterns, and highly impaired functional status. In addition, many older adults during the pandemic have experienced exacerbated social isolation and feelings of loneliness due to disrupted in-person social activities.^21^ The negative consequences of disrupted social and family connectedness, which include death in worst case scenarios,^19,20^ may be particularly salient to nursing home residents, and may underlie the finding of this study that stronger state COVID-19 restrictions were associated with heightened risk of non-COVID-19 mortality among nursing home residents. This finding suggests that, going forward, state public health experts and officials should weight the health benefits of more restrictive social distancing orders (eg, reduced virus transmission) against the negative health consequences due to sustained social disconnectedness. Although the exact balance is largely unknown to us, the recent availability of coronavirus vaccines may make feasible less restrictive shelter-in-place rules for nursing home residents during the remainder of the pandemic.

This study has several limitations. First, we were only able to track state social-distancing measures and nursing home COVID-19 and non-COVID-19 outcomes starting in June due to the lack of national data in early period of the pandemic. Second, although we demonstrated that in aggregate, stronger state social distancing measures had both positive and negative health consequences on nursing home residents, we cannot determine the effect of individual state policies or orders. Finally, our ability to adjust for nursing home, county, and state covariates may be somewhat limited in multivariable analyses. Therefore, the estimated associations may be partially mediated by unmeasured factors that affect COVID-19 outcomes and non-COVID-19 deaths among nursing home residents.

In conclusion, we found that stronger state social distancing measures were associated with improved COVID-19 outcomes (case and death rates), reduced across-facility disparities in COVID-19 outcomes, but more non-COVID-19 deaths among nursing home residents. More restrictive mitigation rules may lead to both health benefits and unintended health consequences.

## METHODS

### Data and sample

The first source of data is the Nursing Home COVID-19 Public File published and updated by the Centers for Medicare and Medicaid Services (CMS, available at https://data.cms.gov/stories/s/COVID-19-Nursing-Home-Data/bkwz-xpvg). This file contains weekly counts of incident COVID-19 cases and deaths among nursing home residents and staff separately, starting from May 25, as submitted by individual nursing homes through CDC’s National Healthcare Safety Network (NHSN) system (COVID-19 LTC Facility Module). These counts were also reported for the week ending on May 24; however, numbers reported for that week may include both new cases (or deaths) in the week and cases (or deaths) identified before that week, which makes the data inappropriate for analyses due to unknown starting dates of reporting for individual nursing homes.^7^ For subsequent weeks, nursing homes reported only new cases and deaths identified in each reporting week. CMS and CDC performed data quality checks to ensure the accuracy of the reported numbers.

We also obtained data on the stringency of state social distancing measures from wallethub.com, based on 17 state COVID-19 policy metrics (e.g. mandatory face masking in public, reopening of restaurants and bars, and workplace temperature screenings). Each metric was graded, by an expert panel, using a 100-point scale, with higher scores representing fewer restrictions; the panel then determined the weighted average across all metrics for each state and used the overall scores to rank-order all 50 states and the District of Columbia. The overall weighted scores and rank orderings were published and updated every 2 to 3 weeks (https://wallethub.com/edu/states-coronavirus-restrictions/73818) starting from May. We downloaded data of state rankings on COVID-19 restrictions in 5 consecutive updates on the following dates: June 9, June 23, July 7, July 21, and August 11.

We linked these state data to nursing home COVID-19 reports assuming a 1-week lag of state policy effect on new nursing home cases (e.g. state rankings on June 9 linked to nursing home case counts for the week of June 15 to June 21; and state rankings on August 11 linked to nursing home case counts for the week of August 17 to August 23), and a 3-week lag on new deaths (e.g. state rankings on June 9 linked to nursing home death counts for the week of June 29 to July 5; and state rankings on August 11 linked to nursing home death counts for the week of August 31 to September 6). We assumed lagged effects of change in state COVID-19 restrictions because it is estimated that the median incubation period of COVID-19 is 5 days^37^ and the median time from illness to death is 18.5 days.^38^

We then linked this longitudinal database to several other data files including: (1) the CMS Nursing Home Compare (NHC) data files (updated in August of 2020) to obtain important covariates of nursing home organizational, staffing, and quality of care measures;^39^ (2) the LTCFocus file created by the Brown University for additional nursing home characteristics; (3) the Area Healthcare Resource File for key county covariates; and (4) the numbers of lab-confirmed COVID-19 cases and deaths for all counties and states obtained from the national database published by the New York Times (https://github.com/nytimes/covid-19-data). These numbers have been compiled and updated in real time by the Times based on reports from state and local public health agencies.

### Variables

The 4 outcomes of interest included nursing home weekly numbers of: (1) new, laboratory-confirmed COVID-19 cases among residents; (2) new, lab-confirmed COVID-19 cases among staff; (3) new COVID-19 related deaths among residents; and (4) new non– COVID-19 related deaths among residents, for each of the 5 reporting weeks described above.

The key independent variable was state rankings on COVID-19 restrictions, which, for ease of presentation, was dichotomized as 1 for states with high stringency (rankings of 26^th^-51^st^) and 0 for states with low stringency (rankings of 1^st^-25^th^). In sensitivity analyses state rankings were included as a continuous variable and, alternatively, were re-categorized as tertile groups in regression analyses. Another key independent variable for the analyses on across-facility disparities in COVID-19 cases and deaths was percentage of racial/ethnic minority residents (African Americans, Hispanics, Asians or Pacific Islanders, and American Indians or Alaskan Natives) in the nursing home (obtained from the LTCFocus file) which was originally defined using the race and ethnicity information from the Minimum Data Set and Medicare enrollment databases. We categorized nursing homes into quartiles to capture possible nonlinear associations between racial composition and COVID-19 outcomes: nursing homes with low proportions of racial/ethnic minority residents (<2.94%, the 25th percentile), medium proportions (2.94%–11.11%, the median), medium-high proportions (11.11%–29.79%), and high proportions (≥29.79%, the 75th percentile).

The following nursing home covariates that were found important to COVID-19 infections or deaths^2-5,7,8^ were included in regression analyses: number of beds, average daily resident census, ownership status (for-profit, non-profit, or government-owned), chain affiliation, hospital affiliation, percentage of Medicare residents, percentage of Medicaid residents, a facility-level case mix index, average staffing levels (hours per resident day) for registered nurse (RN) and for all nursing staff (including RN, licensed practical/vocational nurse [LPN/LVN], and certified nursing assistant [CNA]) in 2019, and five-star ratings for overall quality of care. RN and other nurse staffing levels were calculated based on daily resident census and CMS’ Payroll-Based Journal system through which nursing homes electronically submit the number of hours that agency and contract staff are paid to work each day.^40^ The five-star ratings aggregate ratings of nursing home quality measures on three domains – deficiency citations assigned during on-site inspections, care processes and outcomes of residents, and nurse staffing to resident ratios – into a rating system of one to five stars, with more stars indicating better quality.^41^ Additional nursing home covariates included the cumulative numbers of COVID-19 cases and COVID-19 and non-COVID-19 deaths among residents before state COVID-19 policies were evaluated, which were calculated for the period from May 25 to 2 days before the strength of the state social distancing measures was ranked.

County-level covariates included cumulative number of COVID-19 confirmed cases, cumulative number of COVID-19 deaths, and county population size. State-level covariates included cumulative number of COVID-19 confirmed cases per 1000 population, cumulative number of COVID-19 deaths per 1000 population, percentage of older population (≥65 years), and percentage of non-white population. All county and state cumulative counts were calculated for the period from May 25 to 2 days before the strength of the state social distancing measures was ranked.

### Statistical analyses

We compared differences in nursing home, county, and state characteristics by strength of state COVID-19 restrictions (high vs low) and by nursing home quartile groups of racial/ethnic compositions. T-tests or analyses of variance for continuous variables and chi-square tests for categorical variables were used for statistical inference.

In multivariable analyses we fit separate longitudinal two-part models, with unit of analysis being the nursing home week, to account for the fact that a large number of nursing homes had zero cases (or deaths) in each study week.^42^ The first part of the models was a generalized linear model with a logit link function and an assumed binomial distribution, which estimated the likelihood of a nursing home to have at least one new confirmed case (or death) reported in the week. The second part of the two-part models is a count model that assumed negative binomial distribution to account for the over-dispersion of event occurrence and estimated the number of nursing home new cases (or deaths) conditional on at least one new case (or death) confirmed in the reporting week.

Both parts of the models had an indicator for high vs low strength of state COVID-19 restrictions as the independent variable and controlled for the same nursing home, county, and state covariates, as well as a set of indicators for study weeks (secular trend). In estimating the associations of state policy with across-facility disparities in COVID-19 related outcomes, the two-part models further included interaction between the state policy indicator and indicators for medium, medium-high, and high concentrations of racial/ethnic minority residents in nursing homes (the low concentration group serving as the reference group). All coefficient estimates were based on Huber–White robust standard errors to correct for the correlation of nursing home outcome when it was repeatedly observed over 5 weeks.^43^ After model estimation, we obtained the predicted event counts and plotted each predicted count against different levels of state policy stringency and nursing home racial composition groups.

## Data Availability

All data are available on public databases. (1 Covid-19 Nursing Home Data: https://data.cms.gov/stories/s/bkwz-xpvg, 2 The Nursing Home Compare (NHC) data https://data.medicare.gov/data/nursing-home-compare, 3 Long-Term Care Focus data: % Medicare, %Medicaid, %non-Hispanic whites, chain affiliation http://ltcfocus.org/download/request, 4 Covid 19 county cases and deaths https://github.com/nytimes/covid-19-data, 5 AHRF https://data.hrsa.gov/data/download, 6 rankings of state social distancing measures https://wallethub.com/edu/states-coronavirus-restrictions/73818/).

https://data.medicare.gov/data/nursing-home-compare

http://ltcfocus.org/download/request

https://github.com/nytimes/covid-19-data

https://data.hrsa.gov/data/download

https://wallethub.com/edu/states-coronavirus-restrictions/73818/

## Author Contributions

Li, Cheng, Cai, Mao, Temkin-Greener: conception and design, acquisition of data, analysis and interpretation of data, critical revision for important intellectual content, and final approval of the version to be published. Li: drafting of article.

## Supplements

**Supplementary Table S1.**
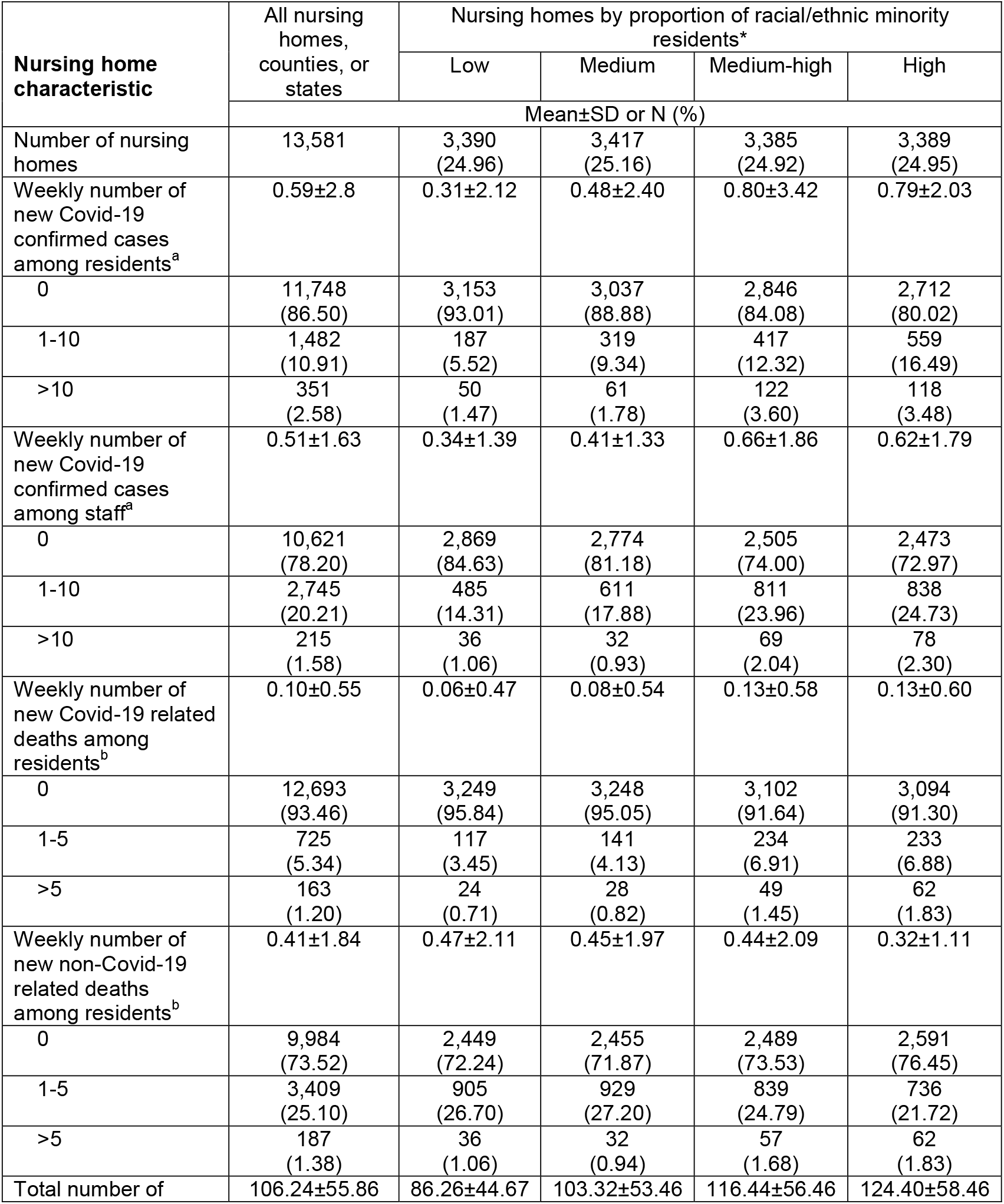
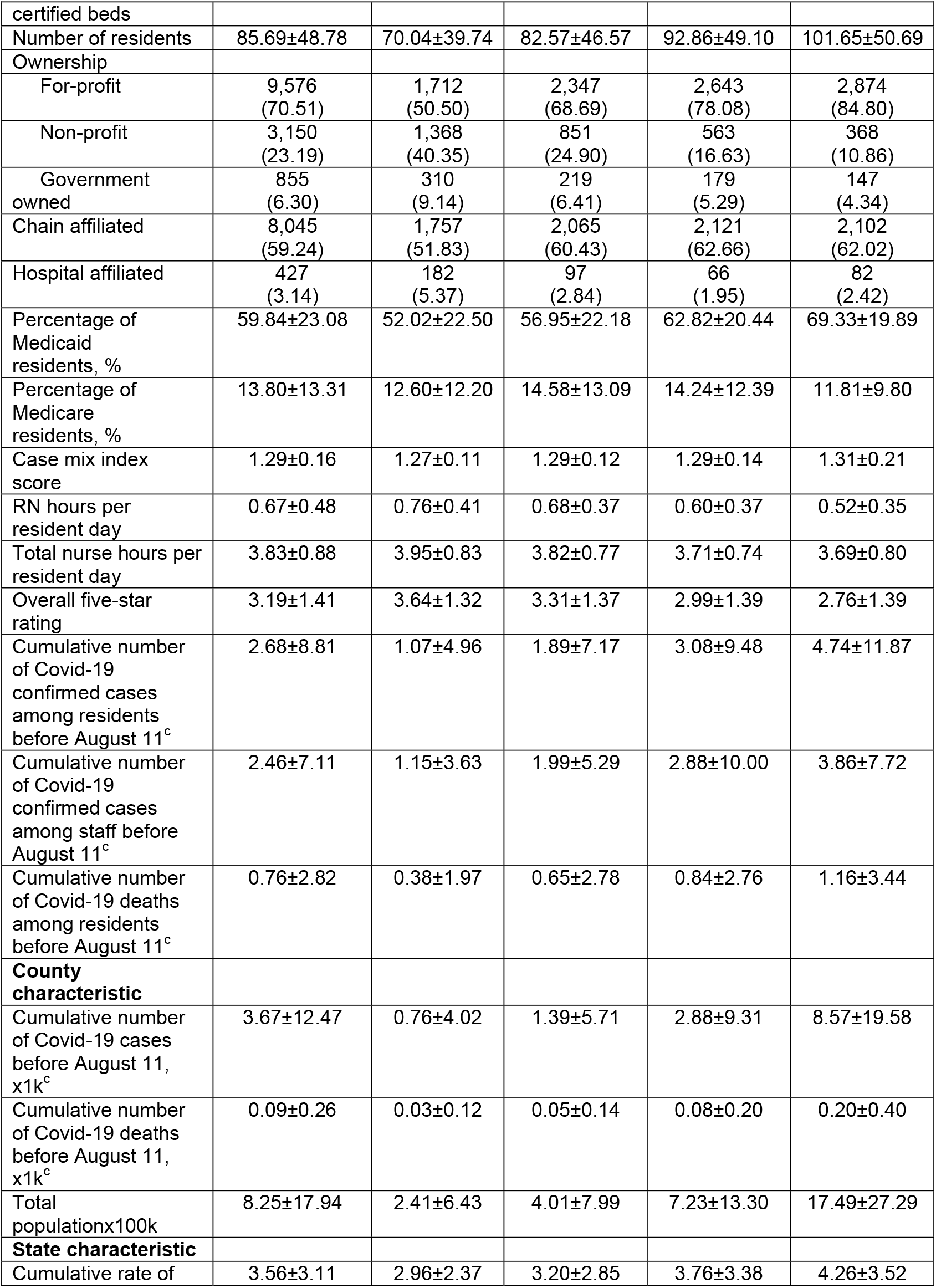
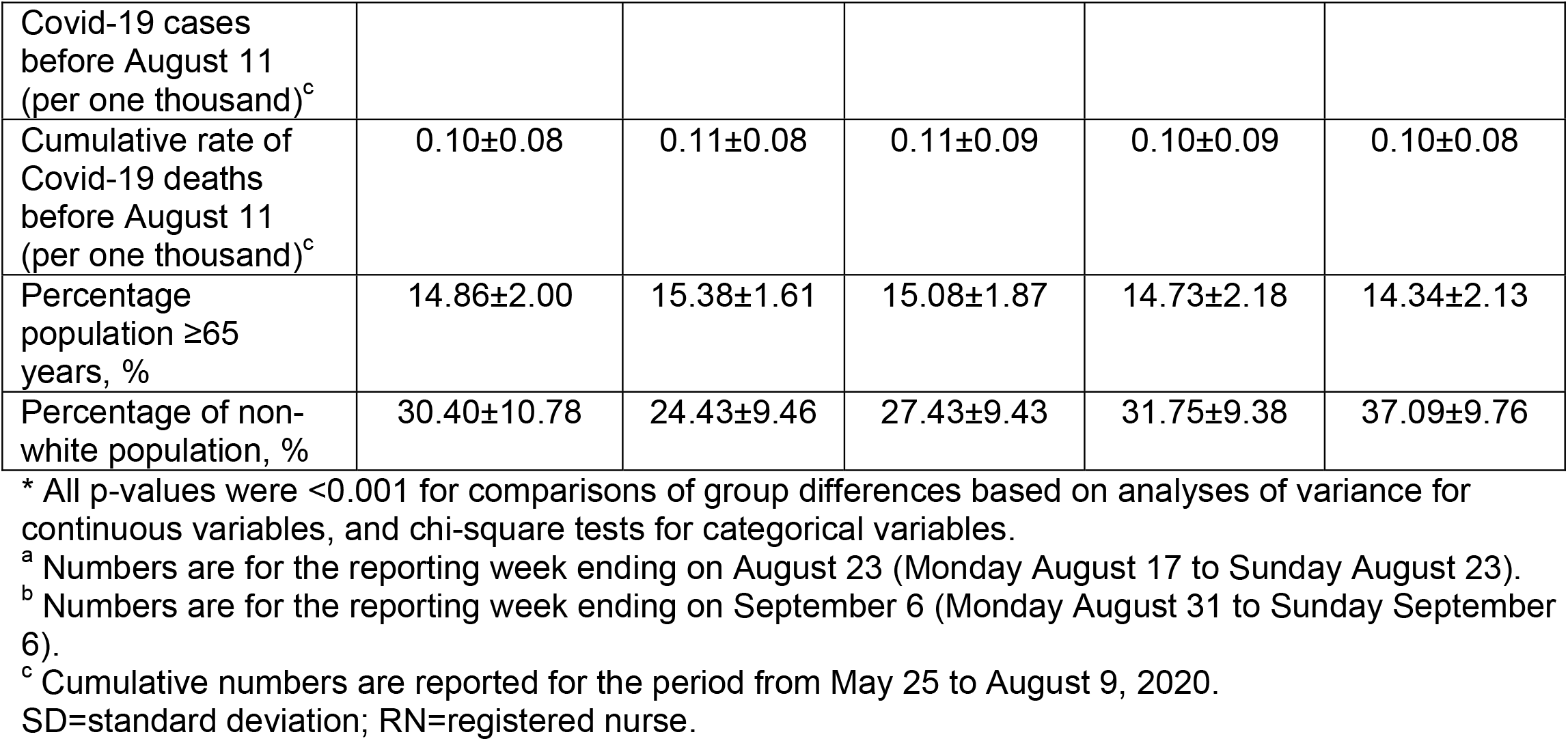
Nursing home characteristics by nursing home quartile groups of proportion of racial/ethnic minority residents.

**Table S2.** Associations between the strength of state social distancing measures and nursing home COVID-19 and non-COVID-19 outcomes – sensitivity analyses in which rankings of state social distancing measures are a continuous variable.^a^.

| **Two-part model for confirmed COVID-19 cases among residents** | **β-coefficient** | **OR or IRR<sup>b</sup>**<br>**(95% CI)** | **P-value** |
| --- | --- | --- | --- |
| **Part 1: likelihood of ≥1 case** |  |  |  |
| Ranking of state social distancing measures x 10 | -0.06 | 0.94 (0.92-0.96) | <0.001 |
| **Part 2: count of cases conditional on ≥1 case** |  |  |  |
| Ranking of state social distancing measures x 10 | -0.04 | 0.96 (0.94-0.99) | <0.001 |
| **Two-part model for confirmed COVID-19 cases among staff** |  |  |  |
| **Part 1: likelihood of ≥1 case** |  |  |  |
| Ranking of state social distancing measures x 10 | -0.08 | 0.92 (0.90-0.94) | <0.001 |
| **Part 2: count of cases conditional on ≥1 case** |  |  |  |
| Ranking of state social distancing measures x 10 | -0.04 | 0.96 (0.94-0.97) | <0.001 |
| **Two-part model for COVID-19 related deaths among residents** |  |  |  |
| **Part 1: likelihood of ≥1 death** |  |  |  |
| Ranking of state social distancing measures x 10 | -0.13 | 0.88 (0.85-0.91) | <0.001 |
| **Part 2: count of deaths conditional on ≥1 death** |  |  |  |
| Ranking of state social distancing measures x 10 | 0.01 | 1.01 (0.99-1.03) | 0.488 |
| **Two-part model for non-COVID-19 deaths among residents** |  |  |  |
| **Part 1: likelihood of ≥1 death** |  |  |  |
| Ranking of state social distancing measures x 10 | -0.003 | 1.00 (0.98-1.01) | 0.706 |
| **Part 2: count of deaths conditional on ≥1 death** |  |  |  |
| Ranking of state social distancing measures x 10 | 0.01 | 1.01 (0.98-1.05) | 0.484 |

<sup>a</sup> Based on two-part models for confirmed cases and deaths separately that adjusted for nursing home, county, and state covariates, time trend, and the clustering of repeated observations of nursing homes.
<sup>b</sup> ORs are reported for part 1 of the two-part models and IRR are reported for part 2 of the two-part models.
OR=odds ratio; IRR=incidence rate ratio; 95% CI=95% confidence interval.

**Table S3.** Associations between the strength of state social distancing measures and nursing home COVID-19 and non-COVID-19 outcomes – sensitivity analyses in which rankings of state social distancing measures are categorized as tertile groups.^a^.

| **Two-part model for confirmed COVID-19 cases among residents** | **β-coefficient** | **OR or IRR<sup>b</sup>**<br>**(95% CI)** | **P-value** |
| --- | --- | --- | --- |
| **Part 1: likelihood of ≥1 case** |  |  |  |
| 2<sup>nd</sup> tertile in strength of state social distancing measures<sup>c</sup> | -0.07 | 0.94 (0.87-1.01) | 0.091 |
| 3<sup>rd</sup> tertile in strength of state social distancing measures<sup>c</sup> | -0.18 | 0.84 (0.78-0.91) | <0.001 |
| **Part 2: count of cases conditional on ≥1 case** |  |  |  |
| 2<sup>nd</sup> tertile in strength of state social distancing measures<sup>c</sup> | -0.10 | 0.90 (0.83-0.99) | 0.022 |
| 3<sup>rd</sup> tertile in strength of state social distancing measures<sup>c</sup> | -0.13 | 0.88 (0.81-0.96) | 0.003 |
| **Two-part model for confirmed COVID-19 cases among staff** |  |  |  |
| **Part 1: likelihood of ≥1 case** |  |  |  |
| 2<sup>nd</sup> tertile in strength of state social distancing measures<sup>c</sup> | -0.07 | 0.93 (0.88-0.99) | 0.015 |
| 3<sup>rd</sup> tertile in strength of state social distancing measures<sup>c</sup> | -0.27 | 0.76 (0.72-0.81) | <0.001 |
| **Part 2: count of cases conditional on ≥1 case** |  |  |  |
| 2<sup>nd</sup> tertile in strength of state social distancing measures<sup>c</sup> | -0.02 | 0.98 (0.92-1.05) | 0.645 |
| 3<sup>rd</sup> tertile in strength of state social distancing measures<sup>c</sup> | -0.14 | 0.87 (0.82-0.93) | <0.001 |
| **Two-part model for COVID-19 related deaths among residents** |  |  |  |
| **Part 1: likelihood of ≥1 death** |  |  |  |
| 2<sup>nd</sup> tertile in strength of state social distancing measures<sup>c</sup> | -0.14 | 0.87 (0.78-0.96) | 0.008 |
| 3<sup>rd</sup> tertile in strength of state social distancing measures<sup>c</sup> | -0.30 | 0.74 (0.66-0.83) | <0.001 |
| **Part 2: count of deaths conditional on ≥1 death** |  |  |  |
| 2<sup>nd</sup> tertile in strength of state social distancing measures<sup>c</sup> | 0.03 | 1.03 (0.95-1.12) | 0.424 |
| 3<sup>rd</sup> tertile in strength of state social distancing measures<sup>c</sup> | 0.04 | 1.04 (0.96-1.12) | 0.331 |
| **Two-part model for non-COVID-19 deaths among residents** |  |  |  |
| **Part 1: likelihood of ≥1 death** |  |  |  |
| 2<sup>nd</sup> tertile in strength of state social distancing measures<sup>c</sup> | -0.09 | 0.91 (0.86-0.97) | 0.003 |
| 3<sup>rd</sup> tertile in strength of state social distancing measures<sup>c</sup> | -0.05 | 0.95 (0.90-1.02) | 0.139 |
| **Part 2: count of deaths conditional on ≥1 death** |  |  |  |
| 2<sup>nd</sup> tertile in strength of state social distancing measures<sup>c</sup> | 0.06 | 1.06 (0.95-1.20) | 0.293 |
| 3<sup>rd</sup> tertile in strength of state social distancing measures<sup>c</sup> | 0.09 | 1.10 (0.98-1.23) | 0.105 |

<sup>a</sup> Based on two-part models for confirmed cases and deaths separately that adjusted for nursing home, county, and state covariates, time trend, and the clustering of repeated observations of nursing homes.
<sup>b</sup> ORs are reported for part 1 of the two-part models and IRR are reported for part 2 of the two-part models.
<sup>c</sup> Compared to the 1<sup>st</sup> tertile group.
OR=odds ratio; IRR=incidence rate ratio; 95% CI=95% confidence interval.

## REFERENCE

1. McMichael TM, Currie DW, Clark S, et al. Epidemiology of Covid-19 in a Long-Term Care Facility in King County, Washington. N Engl J Med. 2020;382(21):2005–2011.

2. Li Y, Temkin-Greener H, Shan G, Cai X. COVID-19 Infections and Deaths among Connecticut Nursing Home Residents: Facility Correlates. J Am Geriatr Soc. 2020;68(9):1899–1906.

3. Harrington C, Ross L, Chapman S, Halifax E, Spurlock B, Bakerjian D. Nurse Staffing and Coronavirus Infections in California Nursing Homes. Policy Polit Nurs Pract. 2020;21(3):174–186.

4. Temkin-Greener H, Guo W, Mao Y, Cai X, Li Y. COVID-19 Pandemic in Assisted Living Communities: Results from Seven States. J Am Geriatr Soc. 2020;68(12):2727–2734.

5. Ouslander JG, Grabowski DC. COVID-19 in Nursing Homes: Calming the Perfect Storm. J Am Geriatr Soc. 2020;68(10):2153–2162.

6. KFF. Kaiser Family Foundation. State Data and Policy Actions to Address Coronavirus. Available at https://www.kff.org/health-costs/issue-brief/state-data-and-policy-actions-to-address-coronavirus/. Dec 23, 2020.

7. Li Y, Cen X, Cai X, Temkin-Greener H. Racial and Ethnic Disparities in COVID-19 Infections and Deaths Across U.S. Nursing Homes. J Am Geriatr Soc. 2020;68(11):2454–2461.

8. He M, Li Y, Fang F. Is There a Link between Nursing Home Reported Quality and COVID-19 Cases? Evidence from California Skilled Nursing Facilities. J Am Med Dir Assoc. 2020;21(7):905–908.

9. Azar KMJ, Shen Z, Romanelli RJ, et al. Disparities In Outcomes Among COVID-19 Patients In A Large Health Care System In California. Health Aff (Millwood). 2020;39(7):1253–1262.

10. Hooper MW, Nápoles AM, Pérez-Stable EJ. COVID-19 and Racial/Ethnic Disparities. JAMA. 2020;323(24):2466–2467.

11. Wadhera RK, Wadhera P, Gaba P, et al. Variation in COVID-19 Hospitalizations and Deaths Across New York City Boroughs. JAMA. 2020;323(21):2192–2195.

12. Rubin-Miller L, Alban C, Artiga S, Sullivan S. COVID-19 Racial Disparities in Testing, Infection, Hospitalization, and Death: Analysis of Epic Patient Data. Published: Sep 16, 2020. Available at https://www.kff.org/coronavirus-covid-19/issue-brief/covid-19-racial-disparities-testing-infection-hospitalization-death-analysis-epic-patient-data/.

13. Smith DB, Feng Z, Fennell ML, Zinn JS, Mor V. Separate and unequal: racial segregation and disparities in quality across U.S. nursing homes. Health Aff (Millwood). 2007;26(5):1448–1458.

14. Weinberger DM, Chen J, Cohen T, et al. Estimation of Excess Deaths Associated With the COVID-19 Pandemic in the United States, March to May 2020. JAMA internal medicine. 2020;180(10):1336–1344.

15. Woolf SH, Chapman DA, Sabo RT, Weinberger DM, Hill L, Taylor DDH. Excess Deaths From COVID-19 and Other Causes, March-July 2020. JAMA. 2020;324(15):1562–1564.

16. Lange SJ, Ritchey MD, Goodman AB, et al. Potential Indirect Effects of the COVID-19 Pandemic on Use of Emergency Departments for Acute Life-Threatening Conditions - United States, January-May 2020. MMWR Morb Mortal Wkly Rep. 2020;69(25):795–800.

17. Czeisler ME, Marynak K, Clarke KEN, et al. Delay or Avoidance of Medical Care Because of COVID-19-Related Concerns - United States, June 2020. MMWR Morb Mortal Wkly Rep. 2020;69(36):1250–1257.

18. Gerst-Emerson K, Jayawardhana J. Loneliness as a public health issue: the impact of loneliness on health care utilization among older adults. Am J Public Health. 2015;105(5):1013–1019.

19. Holt-Lunstad J, Smith TB, Baker M, Harris T, Stephenson D. Loneliness and social isolation as risk factors for mortality: a meta-analytic review. Perspect Psychol Sci. 2015;10(2):227–237.

20. Hoogendijk EO, Smit AP, van Dam C, et al. Frailty Combined with Loneliness or Social Isolation: An Elevated Risk for Mortality in Later Life. J Am Geriatr Soc. 2020;68(11):2587–2593.

21. Kotwal AA, Holt-Lunstad J, Newmark RL, et al. Social Isolation and Loneliness Among San Francisco Bay Area Older Adults During the COVID-19 Shelter-in-Place Orders. J Am Geriatr Soc. 2020.

22. Hsiang S, Allen D, Annan-Phan S, et al. The effect of large-scale anti-contagion policies on the COVID-19 pandemic. Nature. 2020;584(7820):262–267.

23. Courtemanche C, Garuccio J, Le A, Pinkston J, Yelowitz A. Strong Social Distancing Measures In The United States Reduced The COVID-19 Growth Rate. Health Aff (Millwood). 2020;39(7):1237–1246.

24. Lyu W, Wehby GL. Shelter-In-Place Orders Reduced COVID-19 Mortality And Reduced The Rate Of Growth In Hospitalizations. Health Aff (Millwood). 2020;39(9):1615–1623.

25. Padalabalanarayanan S, Hanumanthu VS, Sen B. Association of State Stay-at-Home Orders and State-Level African American Population With COVID-19 Case Rates. JAMA Netw Open. 2020;3(10):e2026010.

26. Liu Y, Mattke S. Association between state stay-at-home orders and risk reduction behaviors and mental distress amid the COVID-19 pandemic. Prev Med. 2020;141:106299.

27. CMS. Center for Clinical Standards and Quality/Quality, Safety & Oversight Group. Suspension of Survey Activities. March 4, 2020. Available at https://www.cms.gov/files/document/qso-20-12-allpdf.pdf-1.

28. CMS. Center for Clinical Standards and Quality/Quality, Safety & Oversight Group. Guidance for Infection Control and Prevention of Coronavirus Disease 2019 (COVID-19) in Nursing Homes (REVISED). March 13, 2020. Available at https://www.cms.gov/files/document/3-13-2020-nursing-home-guidance-covid-19.pdf.

29. CDC. Centers for Disease Control and Prevention, Coronavirus Disease 2019: Preparing for COVID-19: Long-term Care Facilities, Nursing Homes. Available at https://www.cdc.gov/coronavirus/2019-ncov/hcp/long-term-care.html?CDC_AA_refValhttps%3A%2F%2Fwww.cdc.gov%2Fcoronavirus%2F2019-ncov%2Fhealthcare-facilities%2Fprevent-spread-in-long-term-care-facilities.html.

30. CMS. Centers for Medicare and Medicaid Services. COVID-19 Long-Term Care Facility Guidance, April 2, 2020. Available at https://www.cms.gov/files/document/4220-covid-19-long-term-care-facility-guidance.pdf.

31. CMS. Center for Clinical Standards and Quality/Quality, Safety & OversightGroup. Upcoming Requirements for Notification of Confirmed COVID-19 (or COVID19 Persons under Investigation) Among Residents and Staff in Nursing Homes. April 19, 2020. Available at https://www.cms.gov/files/document/qso-20-26-nh.pdf.

32. Li Y, Yin J, Cai X, Temkin-Greener J, Mukamel DB. Association of race and sites of care with pressure ulcers in high-risk nursing home residents. JAMA. 2011;306(2):179–186.

33. Li Y, Mukamel DB. Racial disparities in receipt of influenza and pneumococcus vaccinations among US nursing-home residents. Am J Public Health. 2010;100 Suppl 1:S256–262.

34. Moss K, Wexler A, Dawson L, Long M, Kates J. The Coronavirus Aid, Relief, and Economic Security Act: Summary of Key Health Provisions. Available at https://www.kff.org/coronavirus-covid-19/issue-brief/the-coronavirus-aid-relief-and-economic-security-act-summary-of-key-health-provisions/. April 9, 2020.

35. Berklan JM. Feds increase COVID-19 funding dedicated to nursing homes to nearly $10 billion. Available at https://www.mcknights.com/news/breaking-feds-increase-covid-19-funding-dedicated-to-nursing-homes-to-nearly-10-billion/. July 22, 2020.

36. Dowling MK, Kelly RL. Policy Solutions for Reversing the Color-blind Public Health Response to COVID-19 in the US. JAMA. 2020;324(3):229–230.

37. Lauer SA, Grantz KH, Bi Q, et al. The Incubation Period of Coronavirus Disease 2019 (COVID-19) From Publicly Reported Confirmed Cases: Estimation and Application. Ann Intern Med. 2020;172(9):577–582.

38. Zhou F, Yu T, Du R, et al. Clinical course and risk factors for mortality of adult inpatients with COVID-19 in Wuhan, China: a retrospective cohort study. Lancet. 2020;395(10229):1054–1062.

39. Centers for Medicare and Medicaid Services. Nursing Home Compare, available at https://www.medicare.gov/nursinghomecompare/Data/About.html.

40. Geng F, Stevenson DG, Grabowski DC. Daily Nursing Home Staffing Levels Highly Variable, Often Below CMS Expectations. Health Aff (Millwood). 2019;38(7):1095–1100.

41. CMS. Centers for Medicare and Medicaid Services. Design for Nursing Home Compare Five-Star Quality Rating System, Technical users’ guide. April 2018. Available at https://www.cms.gov/Medicare/Provider-Enrollment-and-Certification/CertificationandComplianc/downloads/usersguide.pdf.

42. Cameron AC, Trivedi PK. Regression Analysis of Count Data. 2nd ed. Cambridge: Cambridge University Press. 2013.

43. White H. A heteroskedasticity-consistent covariance matrix estimator and a direct test for heteroskedasticity. Econometrica. 1980;48:817–830.

